# Protocol for a mixed methods exploratory study of success factors to escalation of care: the SUFFICE study

**DOI:** 10.1101/2021.11.01.21264875

**Authors:** J Ede, P Watkinson, R Endacott

**Affiliations:** Oxford University Hospital NHS Foundation Trust, Oxford, United Kingdom; University of Oxford, Nuffield Department of Clinical Neurosciences, Oxford, United Kingdom; School of Nursing and Midwifery, University of Plymouth, Plymouth, United Kingdom; School of Nursing and Midwifery, Monash University, Melbourne, Australia

**Keywords:** Escalation of care, continuous quality improvement, critical care, decision making, human factors, medical education

## Abstract

**Background:** In the United Kingdom, hospital patients suffer preventable deaths (*failure to rescue*) and delayed admission to the Intensive Care Unit because of poor illness recognition. This problem has consistently been identified in care reviews. Strategies to improve deteriorating ward patient care, such as early warning systems and specialist care teams (Critical Care Outreach or Rapid Response), have not reliably demonstrated reductions to patient deaths. Current research focuses on *failure to rescue*, but further reductions to patient deaths are possible, by examining care of unwell hospital patients who are rescued (successfully treated). Our primary objective is to develop a framework of care escalation success *factors* that can be developed into a complex intervention to reduce patient mortality and unnecessary admissions to the Intensive Care Unit (ICU).

**Methods and Analysis:** SUFFICE is a multicentre mixed-methods, exploratory sequential study examining rescue events in the acutely unwell ward patient in two National Health Service Trusts with Teaching Hospital status. The study will constitute four key phases. Firstly, we will observe ward care escalation events to generate a theoretical understanding of the process of rescue. Secondly, we will review care records from unwell ward patients in whom an ICU admission was avoided to identify care success factors. Thirdly, we will conduct staff interviews with expert doctors, nurses, and Allied Health Professionals to identify how rescue is achieved and further explore care escalation *success factors* identified in the first two study phases. The final phase involves integrating the study data to generate the theoretical basis for the framework of care escalation success factors.

**Ethics and Dissemination:** Ethical approval has been obtained through the Queen Square London Research and Ethics committee (REC Ref 20/HRA/3828; CAG-20CAG0106). Study results will be of interest to critical care, nursing and medical professions and results will be disseminated at national and international conferences.

**Trial Registration Number:** ISRCTN 38850

## Introduction

It has been estimated that there are up to 11,000 preventable deaths in England National Healthcare Services (NHS) Trusts each year (1). Patient deaths, resulting from reversible in-hospital complications, are classified as a Failure to Rescue (FTR) (2,3). FTR is a common theme to National Confidential Enquiry into Patient Outcomes and Death (NCEPOD) reports(4–6). Evidence suggests complication rates do not differ significantly between hospitals, but some patients can be 3 times more likely to die in certain Trusts compared to others (7), implying organisational differences accounting for mortality rates.

## Background

The appropriate use of Intensive Care Unit (ICU) resources is high on the NHS agenda (8). During the Covid-19 pandemic a key focus of the NHS was the optimisation of vital critical care capacity to provide for the surge of patients who would ultimately need advanced level care in a High Dependency Unit (HDU) or ICU (9). Optimisation has two key elements: reducing unnecessary ICU admissions and facilitating timely admissions. Poor recognition of patient deterioration increases patient mortality and morbidity (10) by delaying admission to the ICU. Delays cause worsening patient outcomes (11) including longer ICU and hospital length of stays (12). In order to avoid FTR and maximise ICU treatment efficacy, a successful and timely escalation of care is required (13). Timeliness of a patients’ deterioration detection, communication and management (4,13) (escalation of care) can be influenced by organisational or human factors (system influences affecting human performance). Organisational factors include hospital volume (number of similar procedures completed such as oesophagostomy surgery) (14), nursing ratios, number of critical care beds and the number of operating theatres (15). Human factors attributed to influencing FTR/care escalation include situational awareness, poor clinical monitoring, team working, communication, safety culture, workload, clinical experience, negative emotions and leadership (4,16–20).

Interventions to reduce FTR by optimising care escalation include specialist teams and early warning scores (EWS) (13). Specialist clinical teams, such as Critical Care Outreach or Rapid Response Teams, target improvements to the initial detection and ward management of patient deterioration (21). Unfortunately, evaluations of this intervention conclude a lack of strong evidence of improved outcomes (21,22), such as cardiac arrest rates. Similarly, a recent systematic review on EWS, showed that many tools have methodological weaknesses and may not perform as well as expected (23). Key weakness in EWS, are that they don’t entirely complement the ways in which clinicians escalate care, for example EWS do not integrate soft clinical signals of deterioration (24) often used to escalate a patients care prior to significant illness. Despite these targeted interventions, preventable patient deaths remains problematic (6).

Patient deterioration literature has primarily focused on failure. In the deteriorating ward patient, further reductions to patient mortality and improvements to patient morbidity may be seen by examining the process of rescue and identifying *success factors* to this. SUFFICE forms part of a full-time doctoral study and aims to systematically address gaps in understanding about the process of rescue.

## METHODS

### Aims and Objectives

Our primary aim is to develop a framework of care escalation success factors that can be developed into a complex intervention to reduce patient mortality and unnecessary ICU admissions. Rescue event analysis will be achieved through five key objectives:

- To observe between 200-400 patient escalation events in representative areas
- To review between 200-400 care record reviews of deteriorating patients who avoided an ICU admission
- To interview 30 expert clinical staff to identify success factors and how to apply them effectively
- To describe the patient population in whom escalations occur
- To describe rescue events in Covid-19 negative and positive patients

### Patient and Public involvement (PPI)

This study has been developed and supported by several Patient and Public Involvement (PPI) representatives (CT, IT, MC) that form the SUFFICE PPI group. This group has been consulted at major study milestones including study development and have fed back on study protocol, study design and ethics applications. These representatives are both patient and public, who give a rich insight into the patient experience.

### Study Design

SUFFICE (Success Factors Facilitating Care during Escalation) is a multicentre, mixed-methods, exploratory sequential study aiming to identify the success factors to care escalation (rescue), resulting in a framework development. There are four key study phases; observations of escalation events, a retrospective care records review of ward patients who became unwell but did not require an ICU admission, clinical staff interviews and a data integration phase (see Figure 1. SUFFICE study design flow diagram).

**Figure 1.**
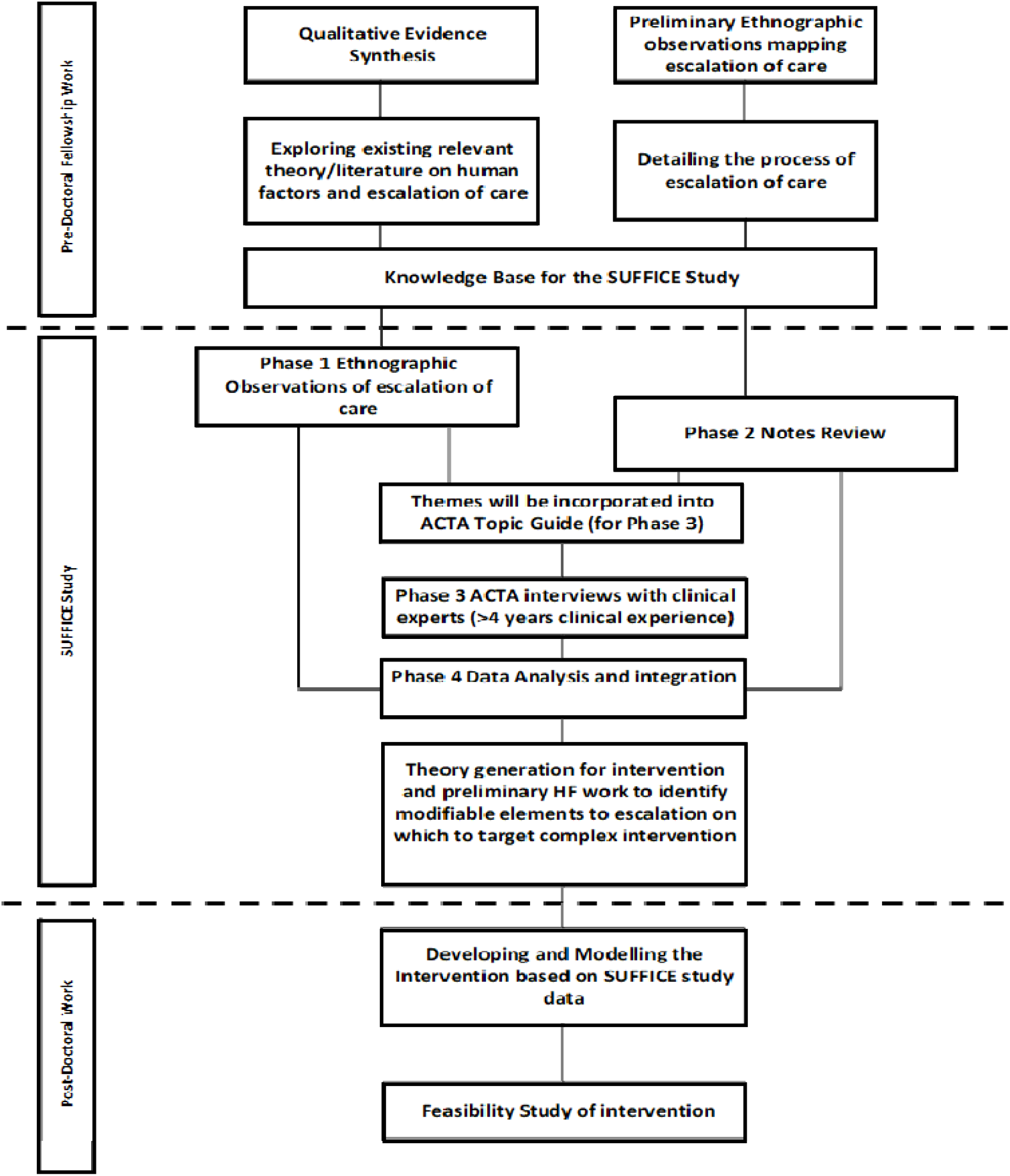
SUFFICE study design flow diagram

### Phase 1: Escalation event observations

Between 200-400 escalation events will be observed to develop a theoretical understanding of the process of rescue. We are defining an escalation of care as any communication relating to the recognition of patient deterioration (13). Non-participant observations (25) will be conducted by the researcher in the clinical ward environment. For the purposes of SUFFICE, observation rather than Ethnography was chosen to ensure an appropriate representation of data collection, which is likely to be short periods of intense fieldwork (26). This will be achieved by shadowing clinical staff and observing their interactions with other staff groups ensuring the collaborative process of rescue is captured. Observations will be conducted in 4-hour sessions within clinical areas, and participants may be observed during multiple observation sessions.

Informal interviews will supplement observations, probing events, staffing levels, or actions, this being an accepted method in observation research (27). Data collected will include patient factors (age, admission type), escalation factors (escalation triggers, EWS), and contextual ward factors (staffing levels). Shelford Safer Care Nursing Tool (SNCT) data, giving an indication of ward staffing levels and ward acuity or dependency (28), may be collected for wards where an escalation event is witnessed. This data collectively will contribute a theoretical understanding of situations that lead to an escalation of care.

### Phase 2: Retrospective Care Records Review (RCRR)

We will review 350 medical, surgical or trauma patient care records to understand why some clinically unwell patients deteriorate to the point where their condition would trigger an intensive care review (a trigger event) but avoid an ICU admission (rescued). We have defined clinically unwell as a EWS score of =>7 which would warrant an ICU review or an admission (29). We have also defined rescue as a resolution of a trigger event without requiring an ICU admission. Care data will be reviewed specifically around periods of care, centred around the patient’s EWS trigger event:

- 24 hours pre-trigger
- 24 hours post-trigger
- >24 hours post-trigger (subsequent care period until 3 subsequent triggers of <3 is documented indicating stability)

A further 50 notes (giving a total of up to 400 care records) will be reviewed from participants who became unwell on the ward were admitted to ICU and died. Records will be reviewed using the Structured Judgement Review (SJR) method (30,31) and will consider all aspects of the patients care by examining records from nursing, Allied Health Professionals, doctors, drugs charts and diagnostic test results (1). Care records reviews are a prime method to assess quality of care (32).

The review process is conducted in three stages: Level 1 and Level 2 (in-depth) reviews:

- Level 1, care records will be reviewed and given quality of care scores before, during and after the trigger event. Quality of care is graded by the reviewer, from 1-5 (1-Very poor care, 2-Poor care, 3-Adequate care, 4 Good care, and 5-Excellent care) in each care period. A small vignette will be documented for each phase justifying rationale of each quality-of-care grade. A modified Case Report Form (CRF) tool will be used to collect the data based on the Structure Judgment Tool (30) used within NHS mortality reviews and has been included within the associated documents. Other data collected will include patient factors; age, length of hospital stay, Clinical Frailty Scale and Charlson Co-morbidity scores, Safer Nursing Care Tool (SNCT) data.
- Level 2 reviews will be conducted on care records that have been graded scores of 4-5 (indicating Good to Excellent care). From these records, a rich qualitative narrative of care factors will be extracted giving a chronology of care pre- and post-trigger. Themes from care reviews may also be explored in Phase 3.
- Validation will consist of a random pragmatic proportion of Level 1 (10%) and Level 2 (n=5) care record reviews being conducted by a second researcher to assess care judgements scores (1-5). A Kappa Coefficient (for interrater reliability) calculated (1) and significant agreement will be assumed with a result of >0.64 (20).The second researcher is likely to be one of the research team (listed as contributors) or a clinical/research colleague with suitable expertise, training and trust clearances.

### Phase 3: Staff Applied Cognitive Task Analysis (ACTA) interviews

We will interview up to 30 nursing and medical experts with at least 4 years’ clinical experience, to understand factors affecting successful care escalation and identify how these could be applied effectively across healthcare settings. This may be staff who also participated in Phase 1. Interviews will be guided by a piloted interview developed using the Applied Cognitive Task Analysis (ACTA) interviewing methodology. ACTA is a cognitive interviewing technique, not requiring specialist cognitive training, and centres on eliciting *expert* knowledge used to perform key tasks (33). Expert participants will describe how they manage patient deterioration and care escalation (34), articulating their knowledge through posed escalation scenarios. The focus of the interviews is to identify escalation success factors, identified by healthcare experts.

The broad ACTA interview schedule is as follows:

**<u>Task diagram</u>**: Asks participants to list six key escalation tasks. Aims to get the interviewee focussed on escalation tasks and creates a process map (ordered diagram of escalation).

**<u>Knowledge Audit</u>**: Identifies how expertise is utilised during escalation. Escalation questions are organised around expertise categories: diagnosing, predicting, situational awareness, perceptual skills, workarounds, improvising, meta cognition and recognizing anomalies.

**<u>Simulation Interview</u>**: Interviewee is posed an escalation simulation prompting expertise which may not have already developed from the knowledge audit.

To maximise opportunities to collect this specific information, an experience cut-off for participant selection has been given. Participant interviews (over the telephone or face to face) will last no longer than 90 minutes (60-90 minutes), will be digitally recorded on an encrypted device and transcribed.

### Phase 4: Data Analysis and Integration

A full study data analysis plan is outlined in the analysis section.

### Setting

This study is taking place in two separate United Kingdom NHS Trusts. These are two contrasting Trusts, one being a large teaching hospital and the other being a smaller DGH. Trust A is a group of three tertiary referral hospitals and one district general hospital. In total, the organisation has approximately 1,465 beds, and serves a population of around 655,000. The intensive care capacity within this trust comprises on three specialist units, Neuro, Cardiac and General. The total Trust ICU bed capacity is approximately 48 beds with no established Critical Care Outreach Team. Trust B is the main provider of acute hospital services for the population of approximately 500,000 people. The hospital has approximately 813 inpatient beds of which 627 are acute bed, 66 for children and young people 75 maternity. This trust has a well-established, nurse-led critical care outreach team and an ICU capacity of 16 beds.

### Participant Selection

#### Phase 1: Escalation event observations

Clinical staff who receive or make escalation referrals will be approached and consented for observations. Staff will be purposively selected to ensure a variety of specialities (surgical, trauma, medical) and clinical grades. Participants should be over the age of 18 and be willing to give informed consent. Observations will cease when a full data set (of 200-400 witnessed escalation events) is completed. Staff will be recruited through posters, word of mouth and invitation emails endorsed by key ward stakeholders.

#### Phase 2: Retrospective Care Records Review (RCRR)

Our focus is to identify acutely unwell medical, surgical or trauma patients in whom a ward rescue event has occurred without needing an ICU admission. We have defined acutely unwell as a EWS =>7 (35). A EWS score of =>7 has a higher specificity, predictive value, positive likelihood ratio and positive predictive value of an ICU admission than a threshold of 5, and therefore these patients have a high probability of an ICU admission (36). The aim of this analysis is to identify why these patients (with a high probability of needing an ICU admission) avoid this event and if and how they are rescued. This trigger value was also supported through study team consensus as this physiological derangement would be unlikely to spontaneously resolve without intervention. Patients who meet the inclusion criteria will be identified from the local electronic patient record (EPR). The most recent patients who meet the inclusion criteria will be identified and their care records retrieved.

### Record reviews survivors

- Medical, surgical or trauma patients who have had an EWS=>7
- Have not been admitted to ICU
- Survived their hospital admission (discharged to normal place of residence)
- Have Covid-19 and non-Covid-19 infections

### Record reviews deceased

- Medical, surgical or trauma patients have triggered a=>7 EWS score
- Have been admitted to ICU
- Died
- Have Covid-19 and non-Covid-19 infections

Records will be sampled from varying timepoints within the year to ensure seasonal variation is captured. If notes are incomplete, these will be excluded until 400 usable datasets are completed. Confidentiality Advisory Group (CAG) (CAG-20CAG0106) support was obtained for this study (see ethics section).

#### Phase 3: Staff Applied Cognitive Task Analysis (ACTA) interviews

Any staff member who has experience of escalating or receiving a care escalation and has 4 years or greater clinical experience are eligible to take part in the interview. To maximise opportunities to elicit expertise utilised during escalation of care, an experience cut-off for participant selection has been given. For the purposes of this study, ‘expert’ is defined as greater than 4 years clinical experience (37,38). Pilot interviews have been conducted exploring care escalation. Participants will be chosen for representativeness in terms of gender, age and clinical role, ensuring varied responses. Staff will be over the age of 18 and are willing to give informed consent. If a staff member withdraws from the study, they will be replaced to ensure a full dataset is collected. Staff will be recruited through posters, word of mouth and invitation emails endorsed by key ward stakeholders.

#### Phase 4: Data Analysis and Integration

n/a

### Sample Size

#### Phase 1: Escalation event observations

Observations will be undertaken in four-hour time slots, at varied times of day but with a maximum of one time slot per day. Data collection will continue for 6-12 months; estimates (from preliminary observation work) are that between 8-10 care escalations will be captured per observation session, generating between 200-400 escalation episodes in total. Saturation of data themes will be used as an indicator of sample size (39).

#### Phase 2: Retrospective Care Records Review (RCRR)

This sample size (400 care record reviews) is a pragmatic choice with each Level 1 review estimated to take 1 hour to complete. Each in-depth Level 2 review may take up to four hours to complete. Based on other studies using this method (31,40) between 200-400 notes were a large enough number that the data will be sufficiently rich, and saturation is likely to be achievable.

#### Phase 3: Staff Applied Cognitive Task Analysis (ACTA) interviews

For qualitative interviewing it is accepted that 12 interviews are likely to generate data saturation (41). We chose not to dilute the number of interviews as participants will be selected from two participating NHS sites. It was felt that 15 interviews from each site would generate a rich data set. Interviews will continue until the full 30, or until data saturation is achieved.

#### Phase 4: Data Analysis and Integration

n/a

### Data storage

All data will be stored as per the General Data Protection Regulations (GDPR). All electronic data will be stored on a secure NHS server that was password protected. All paper documentation will be stored in a locked NHS or research facility and appropriately archived once the study has been completed.

### Data Analysis

#### Phase 1: Escalation event observations

Statistical analysis will be mostly descriptive and may include (but not limited to) patient factors collected using the Charlson Comorbidity Index (CCI) tool and the Clinical Frailty Scale (CFS). For continuous data (includes but not limited to) triggering patient factors, escalation data and contextual organisational factors, mean and standard deviation will be calculated. For categorical data (includes but not limited to) escalation type, organisational data, number and percentage will be reported. This will provide context with which to analyse the qualitative data and identify patterns within and across data collection settings. Statistical advice will be sought for this study.

For qualitative data, a Framework Analysis method was chosen as it provides a clear structured output in the form of a Coding Matrix (42). There are 5 key steps to be taken within a Framework Analysis (43) which include:

- Familiarisation
- Identifying a thematic framework
- Indexing (selecting the interesting fragments-coding)
- Charting/Summarising (key difference between this and content analysis) Tell the story of those fragments
- Interpretation

#### Phase 2: Retrospective Care Records Review (RCRR)

Our primary objective is to identify care escalation success factors. To generate an in-depth understanding of this, hospital factors will be explored against care scores generated through notes reviews. Care records data will be extracted using the SJR forms designed by the Royal College of Physicians (RCP) (44) (see methods for full details). Care record review data collected will consist of ordinal (care scores, CFS) and continuous variables (agency usage, bed occupancy rates, staffing ratios). Continuous variables will be described with means, standard deviations, medians, and inter-quartile ranges. Categorical (ordinal) variables will have number and percentages. If the data obtained do not violate any statistical principles, we may model the association of care scores with other ward-level variables such as agency usage, bed occupancy and staffing levels using an Ordinal Regression. All tests will be performed in SPSS. P-values will be considered statistically significant when less than 0.05.

A Kappa Co-efficient will also be calculated using SPSS to indicate the level of interrater agreement between quality of care judgement scores (1) from both reviewers completing care record reviews. If data indicates poor interrater agreement, joint reviews will be undertaken to ensure consistency of methods and explanations for scores will be documented within a study audit trail.

For qualitative data, a rich narrative of care will be obtained from notes reviews using a piloted data collection form and will be thematically coded (45). This includes duplicate data extraction (by two researchers or a coding buddy) for up to 40 Level 1 care reviews and 5 Level 2 care reviews.

#### Phase 3: Applied Cognitive Task Analysis (ACTA) Interviews

Interview transcript data will be coded using Thematic Analysis (45) and inserted into a Knowledge Audit Table and Cognitive Demands Tables (similar to a framework analysis) using the headings from the original ACTA methodology (33) (see methods section of interview structure). The aim of this table (as with Framework Analysis) is to help the researcher identify common themes or conflicting information. A second researcher will cross-analyse up to 3 interviews to ensure coding rigor and theme identification. However, the coding process will be subject to strict collaboration between the research team. To strengthen rigor and credibility, a coding audit trail will be completed detailing coding decisions. Data will be analysed in a coding software package such as NVivo.

#### Phase 4: Data Analysis and Integration

In this final phase, study data will be analysed, linked and compared (viewed as a whole or mixed) (46) enabling the research team to generate a framework of care escalation success factors. Full study data will be analysed together after initial analysis (in each of the phases), defining areas of data convergence or divergence (47). Following data tabulation, success factors to escalation will be identified (intervention framework). Work will be undertaken by the study team, a Human Factors scientist, and the stakeholder group, using knowledge from SUFFICE and the existing body of escalation literature, to develop this framework.

The framework of success factors may include:

- Success factor (description of the factor)
- Outcome (what outcome that factor facilitates)
- Context (what is the context to that factor such as ward, patient)
- Knowledge base (what is understood about that factor already in the literature)
- Balancing measure (could there be any negative system outcomes if that factor were implemented)

## ETHICS AND DISSEMINATION

### Ethics

This study gained ethical approval from the Queens Square London Research Ethics Committee (HRA-20HRA/3828; CAG-20CAG0106). The Oxford University Hospital Foundation NHS Foundation Trust will act as sponsor. The study has been developed with the support of the SUFFICE PPI group. This paper reports SUFFICE Protocol Version 2.1 and has been written with reference to the SPIRIT (Standard Protocol Items: Recommendations for International Trials) checklist (48) (see Supplementary File 1). This protocol paper has been independently peer reviewed by an expert (CS) within the field of patient deterioration and Critical Care.

### Phase 1 and Phase 3

Informed consent will be obtained from participants by trained researchers. This study has been developed with the help of the Trust’s clinical psychology department and staff wellbeing has been carefully considered. If there is distress caused by participating in this study, then staff will be referred to the Trust’s occupational departments and the study team will again review the methods with a clinical psychologist. This study is about identifying effective patient care, but in very rare circumstances this may identify poor care which may require escalation through the local clinical governance channels.

### Phase 2

CAG (CAG-20CAG0106) supports the study team to screen hospital patient lists (including name, DOB and Hospital number) for eligible patients under Section 251 of the NHS Act 2006. The report of patients meeting the inclusion criteria will be generated by the hospital information team and will not be extraneous to the purpose of the study and only allow for eligibility assessment. From this report, the researcher (ICU nurse and another suitable clinician) will access patient notes to perform the Retrospective Care Records Review (as detailed in methods). The Trust generated data report will be held within an NHS network. Only a researcher with a clinical background will use this report and similarly patient electronic records. Once the record reviews have been conducted, the identifiable data will be destroyed.

### Phase 4

n/a

### Dissemination

Results from this study will be disseminated at regional and international conferences. These conferences will be attended by one of the SUFFICE PPI representatives should they choose. Papers generated will be published in peer reviewed medical and nursing journals.

## Supporting information

SPIRIT Checklist

## Data Availability

All data produced in the present study are available upon reasonable request to the authors

## Contributor statement

JE designed and led the project. RE and PW have substantially contributed to this work, provided expertise and PhD supervision, and have agreed the final manuscript.

## Disclaimer

This paper presents independent research funded by the National Institute for Health Research (NIHR). The views expressed are those of the authors and not necessarily those of the NHS, the NIHR or the Department of Health and Social Care.

